# Impact of Quality Improvement Support on Hospital Readmissions and Patient Safety Outcomes: A Quasi-Experimental Study of a National Quality Improvement Initiative

**DOI:** 10.1101/2025.09.26.25336719

**Authors:** Rohit Borah, Andrea Acevedo, Qingkun Shang, Ashlie Wilbon Gyr, Cassandra Carter, Geoffrey Berryman, Nancy Sonnenfeld

**Author notes:** Corresponding author: Rohit Borah, MPH.

## Abstract

**Background:** The Centers for Medicare & Medicaid Services (CMS) administers the Hospital Quality Improvement Contractor (HQIC) program to improve healthcare quality and safety in hospitals serving Medicare beneficiaries. These contractors provided technical assistance to small, rural hospitals, and/or to hospitals with lower quality ratings. This study aimed to evaluate the program’s effectiveness in addressing two priority areas: reducing hospital readmissions and increasing patient safety.

**Methods:** A quasi-experimental design was used to assess the impact of HQIC support on 30-day all-cause readmissions and eight patient safety outcomes in hospitals that received support between October 1, 2020, and December 31, 2023 (N = 1,929). Patient safety outcomes included adverse drug events (ADEs) and healthcare-associated infections (HAI). A difference-in-difference model specification with a Poisson generalized estimating equation was used to compare hospitals receiving support for each outcome to matched hospitals that did not receive HQIC support.

**Results:** Receiving HQIC support was associated with a 1.4% reduction in the number of 30-day all-cause readmissions compared to hospitals with similar characteristics that did not receive HQIC support for readmissions. Hospitals receiving HQIC support for reducing catheter-associated urinary tract infections (CAUTI) showed a 17.4% reduction in the number of CAUTI events relative to hospitals that did not receive support. There was no statistically significant effect on the other outcomes.

**Conclusions:** The HQIC program support had an impact on reductions in hospital readmissions and reductions for one of the HAIs but not on other patient safety outcomes. These findings suggest a targeted approach in resource allocation may improve readmissions and CAUTI rates for small and rural hospitals.

**Key Messages:** *What is already known on this topic:*

- Hospital readmissions, adverse drug events in inpatient care, and healthcare-associated infections increase morbidity and mortality and are costly to the healthcare system, and evidence-based strategies exist to help reduce these events.
- The Centers for Medicare & Medicaid Services implemented a national quality improvement initiative that provides training and technical assistance to hospitals, particularly to small and rural hospitals, to reduce readmissions and improve patient safety outcomes.

*What this study adds:*

- This study provides the opportunity to examine the impact of a national quality improvement effort on hospital readmissions and patient safety outcomes. Quality improvement support was associated with significant reductions in hospitals’ 30-day all-cause readmissions and catheter-associated urinary tract infections, but not with other patient safety indicators assessed.

*How this study might affect research, practice or policy:*

- The finding that quality improvement support to hospitals resulted in improved outcomes suggests the need for prioritization and investment in this approach.
- The limited program impact on improving rare outcomes highlights the importance of aligning goals with the unique needs of smaller, rural hospitals in the design of future quality improvement programs.

## INTRODUCTION

Despite decades of targeted interventions to improve hospital quality of care in the United States, there remains room for improvement. Nationally, approximately 14% of hospital admissions result in readmission within 30 days of discharge.^1^ It is estimated that 1 in 31 inpatient stays have a healthcare-associated infection (HAI) obtained while receiving care for a different condition.^2^ Hospital readmissions, HAIs, and other patient safety indicators are associated with premature deaths, morbidity and excess costs.^3–8^

Reducing hospital readmissions and improving the quality of care for Medicare beneficiaries are long-standing priorities for the Centers for Medicare & Medicaid Services (CMS).^9–11^ Medicare, a federal health insurance program, covers over 67 million Americans, primarily adults aged 65 and older along with younger individuals with long-term disabilities..^12^ Medicare Part A covers inpatient care and coverage is available at no cost to eligible beneficiaries, while other components, such as those covering outpatient services and prescription drugs, are offered to beneficiaries at an additional cost. The Hospital Quality Improvement Contractor (HQIC) program is a CMS-led initiative targeting improvements in hospital healthcare quality and safety. CMS partners with quality improvement (QI) contractors to collaborate with hospitals and serve as QI experts, facilitators, and change agents.

Unlike broader, national QI programs, the HQIC program was designed to address the unique challenges faced by small and rural hospitals. Providing QI support to these hospitals ensures that all beneficiaries have access to high-quality care, improves outcomes, and enhances healthcare value.

However, the support provided is not targeted to Medicare beneficiaries specifically and benefits all patients served in enrolled hospitals.

In its most recent contract, the HQIC program had three priority areas: improving patient safety, increasing quality of care transitions to reduce hospital readmissions, and decreasing opioid misuse. The HQIC program targeted hospitals that were small, located in rural areas with limited access to high-quality care, served high proportion of patients with low socioeconomic status, and/or hospitals with lower national quality ratings.^13^ Nine organizations were selected as HQIC contractors (HQICs) to support eligible hospitals across 50 states and five territories. HQICs provided support to participating hospitals to improve quality of care and to respond to emerging public health crises.

The objective of this study was to assess the effectiveness of the HQIC program for two priority areas: hospital readmissions and patient safety outcomes. Specifically, we aimed to determine the impact of receiving HQIC support on improving the following among Medicare beneficiaries admitted to HQIC- enrolled hospitals: 1) 30-day all-cause hospital readmissions, 2) hypoglycemia-related adverse drug events (ADEs), 3) anticoagulation ADEs, 4) pressure injuries, 5) postoperative sepsis, and 6) four HAIs: catheter-associated urinary tract infection (CAUTI), central-line-associated bloodstream infection (CLABSI), clostridium difficile (C.diff), and methicillin-resistant staphylococcus aureus (MRSA).

## METHODS

### Hospitals, HQICs and Interventions

CMS identified 2,638 eligible hospitals across the United States. The program targeted rural hospitals with specific attention to critical access hospitals. Critical access hospitals are designated by CMS as small, rural facilities (≤25 beds) located at least 35 miles from another hospital. Hospitals with a history of lower performance, identified as having a quality rating of 2 or lower (out of 5) in national quality ratings,^13^ were also a priority.

Nine HQICs were responsible for recruiting and enrolling eligible hospital. Enrollment began in September 2020, and a total of 1,929 hospitals (73.1%) enrolled by December 31, 2023.

CMS encouraged contractors to work with hospital stakeholders to achieve targets through QI activities using evidence-based practices. HQICs also provided hospitals with support on data collection and analytics to inform efforts. CMS designed the program to give contractors flexibility to tailor support based on each hospital’s specific needs. Factors like rural location, hospital size, critical access status, past performance, and safety concerns such as high rates of ADEs, HAIs, or readmissions were all considered.

For example, hospitals with high rates of C. Diff infections received focused help in the form of infection prevention training, improved antibiotic use, and clinical toolkits. Facilities with ADEs received support on safer prescribing and pain management strategies. Smaller rural hospitals with limited experience in QI received basic training on data collection and root cause analysis and participated in peer learning groups. This targeted support helped ensure hospitals were not overwhelmed with information but instead received support that matched their specific clinical challenges and the reality of their facility.

HQICs implemented a wide range of in-person, virtual, and hybrid intervention activities, including training hospital staff on evidence-based resources such as checklists and toolkits, providing technical assistance with implementation of those practices, holding office hours, and training staff on quality delivery of services. HQICs also provided data-related support, including helping hospitals understand QI data and developing data collection protocols to inform QI activities. Descriptions of the types of intervention activities can be found in Table 1 in the Supplemental Material. HQICs were asked to collect data on the type of support they provided hospitals using a structured template based on outcome area and intervention category and to submit data quarterly.

**Table 1.**
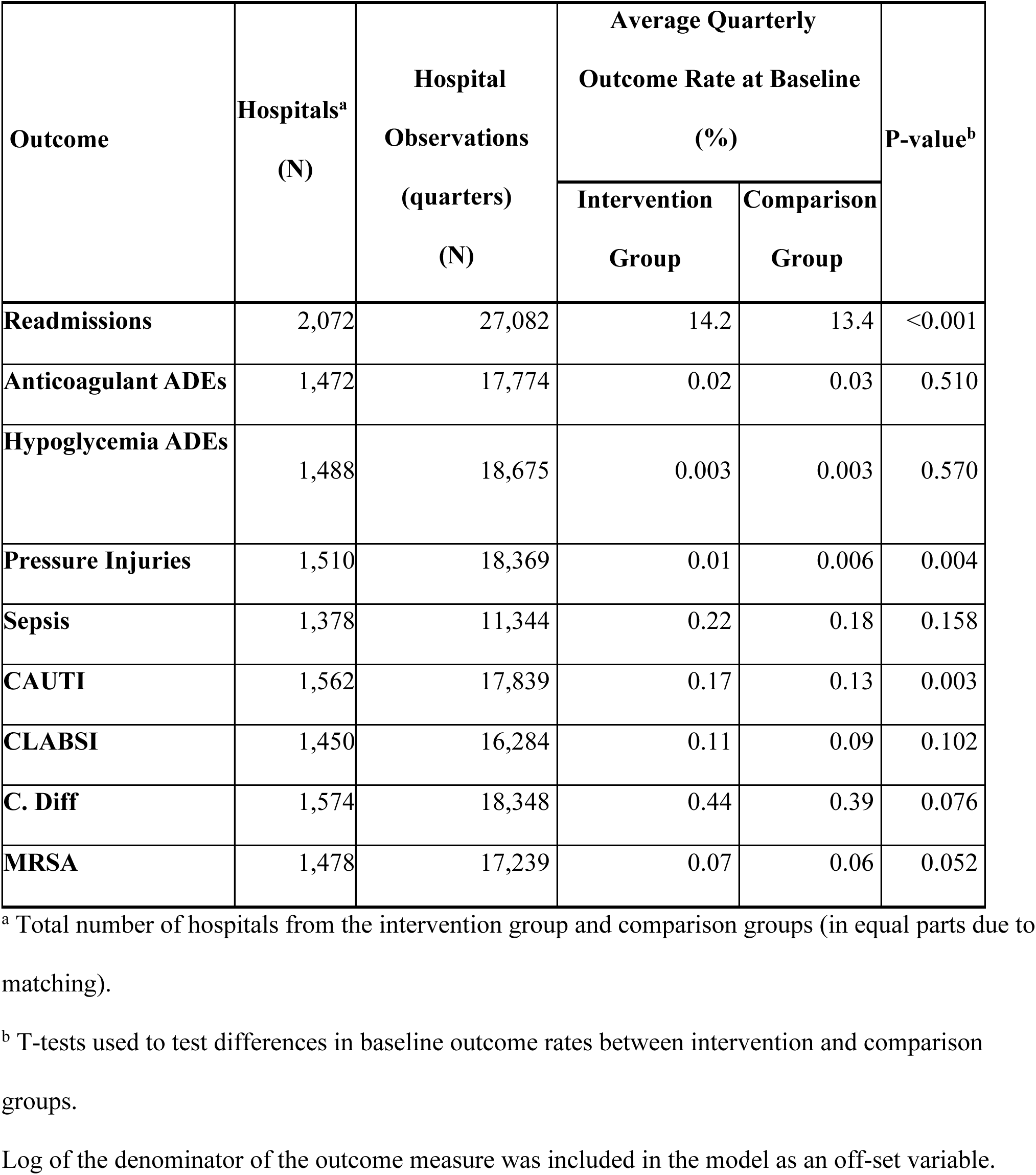
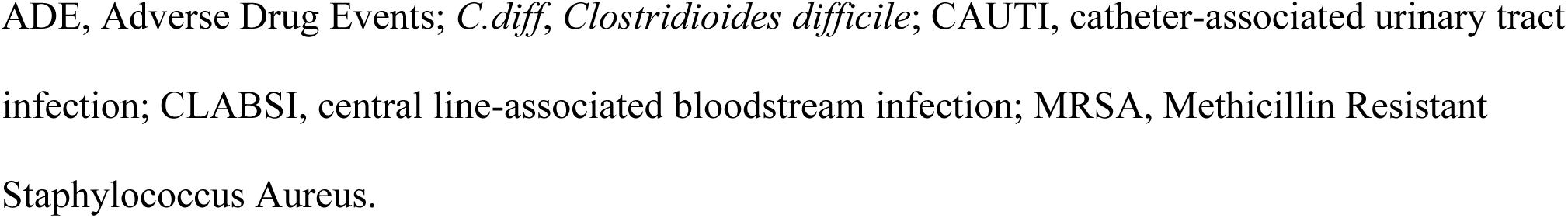
Hospitals and hospital observations included in analytic samples and baseline outcome rates.

| Outcome | Hospitals<sup>a</sup><br>(N) | Hospital<br>Observations<br>(quarters)<br>(N) | Average Quarterly<br>Outcome Rate at Baseline<br>(%) |  | P-value<sup>b</sup> |
| --- | --- | --- | --- | --- | --- |
|  |  |  | Intervention<br>Group | Comparison<br>Group |  |
| **Readmissions** | 2,072 | 27,082 | 14.2 | 13.4 | <0.001 |
| **Anticoagulant ADEs** | 1,472 | 17,774 | 0.02 | 0.03 | 0.510 |
| **Hypoglycemia ADEs** | 1,488 | 18,675 | 0.003 | 0.003 | 0.570 |
| **Pressure Injuries** | 1,510 | 18,369 | 0.01 | 0.006 | 0.004 |
| **Sepsis** | 1,378 | 11,344 | 0.22 | 0.18 | 0.158 |
| **CAUTI** | 1,562 | 17,839 | 0.17 | 0.13 | 0.003 |
| **CLABSI** | 1,450 | 16,284 | 0.11 | 0.09 | 0.102 |
| **C. Diff** | 1,574 | 18,348 | 0.44 | 0.39 | 0.076 |
| **MRSA** | 1,478 | 17,239 | 0.07 | 0.06 | 0.052 |

<sup>a</sup> Total number of hospitals from the intervention group and comparison groups (in equal parts due to matching).
<sup>b</sup> T-tests used to test differences in baseline outcome rates between intervention and comparison groups.
Log of the denominator of the outcome measure was included in the model as an off-set variable.
ADE, Adverse Drug Events; *C.diff*, *Clostridioides difficile*; CAUTI, catheter-associated urinary tract infection; CLABSI, central line-associated bloodstream infection; MRSA, Methicillin Resistant Staphylococcus Aureus.

A dedicated team at CMS provided program oversight and monitoring. The team met with HQICs regularly to interpret QI metrics, provide guidance, and ensure fulfillment of program requirements.

### Design

We used a quasi-experimental design to assess the impact of receiving HQIC support on readmissions and patient safety outcomes in enrolled hospitals through December 31, 2023. The study population included Medicare-certified hospitals with outcome data available through CMS Medicare Fee-for-Service (FFS) Part A claims and Centers for Disease Control and Prevention (CDC) National Healthcare Safety Network (NHSN) HAI data. Rural emergency hospitals without dedicated beds for hospitalizations (N = 11) were excluded.

A quasi-experimental design was used as the facilities were assigned to groups based on specific criteria as opposed to randomized assignment. HQIC-enrolled hospitals did not necessarily receive support for every outcome, and the timing of when each hospital began receiving support for specific outcomes varied. Therefore, intervention and comparison groups were identified for each outcome separately. Intervention groups were composed of enrolled hospitals that received technical assistance from their HQIC for that outcome between October 1, 2020, and December 31, 2023. The comparison group consisted of three subgroups: hospitals that were ineligible to receive HQIC support, eligible hospitals that did not enroll in the HQIC program, and hospitals enrolled in the HQIC program that did not receive HQIC support for that outcome. Figure 1 shows a graphical representation of the criteria used for deciding intervention and comparison groups.

**Figure 1.**
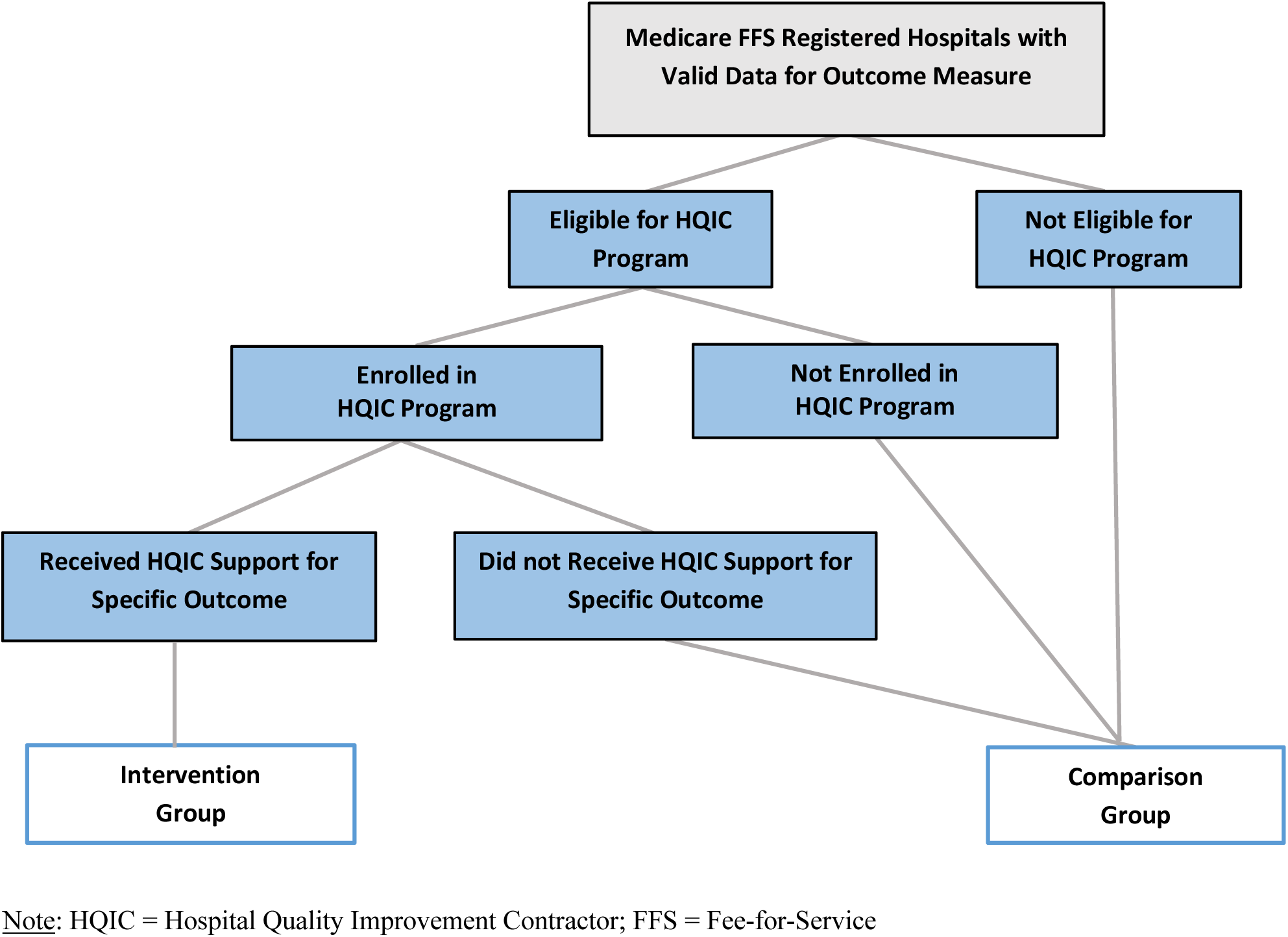
Criteria for intervention and comparison hospital groups.

### Data Sources

The data sources included Medicare Part A FFS claims, NHSN data, and information collected from HQICs on the interventions provided to the enrolled hospitals. Medicare Part A FFS claims were used to create samples, calculate numerators and denominators for most outcome measures, and calculate covariates related to patient demographics. Medicare FFS is a program in which hospitals are reimbursed for services performed as opposed to a bundled payment per beneficiary managed by a private health plan. The NHSN dataset, maintained by the CDC, is a national HAI tracking system and was used to obtain data on HAIs at each hospital. Claims and NHSN data from October 1, 2019, to March 31, 2024, were used in the analysis. This approach allowed for all hospitals to have at least one year of baseline data prior to receipt of HQIC support. Hospitals that began receiving support for a new outcome during late 2023 had at least one quarter of data after starting to receive HQIC support. We also used HQIC-reported data describing their intervention activities provided to each hospital.

HQICs were required to report quarterly on the interventions provided using a structured template that included the type of support provided, the outcome(s) targeted, the hospital receiving the support, and the start date for each specific type of intervention. Other sources were used to obtain information on hospital characteristics used as covariates. Table 2 in the Supplemental Material provides more detailed descriptions of all data sources used.

**Table 2.** Unadjusted and adjusted estimated reduction attributed to receiving HQIC support following initial intervention.

| **Outcome** | **Unadjusted Models** |  |  | **Adjusted Models** |  |  |
| --- | --- | --- | --- | --- | --- | --- |
|  | **Coefficient**<br>**(95% CI)** | **Mean**<br>**Attributable**<br>**Reduction**<br>**(95% CI)** | **P-value** | **Coefficient**<br>**(95% CI)** | **Mean**<br>**Attributable**<br>**Reduction**<br>**(95% CI)** | **P-value** |
| **Readmissions** | -0.01<br>(-0.02, 0.002) | -1.0%<br>(-2.2%, 0.2%) | 0.103 | -0.01<br>(-0.03, -0.002) | -1.4%<br>(-2.5%, -0.1%) | 0.028 |
| **Anticoagulant**<br>**ADEs** | -0.01<br>(-0.19, 0.16) | -1.2%<br>(-17.1%, 17.8%) | 0.892 | -0.03<br>(-0.20, 0.15) | -2.5%<br>(-17.9%, 15.7%) | 0.770 |
| **Hypoglycemia**<br>**ADEs** | 0.04<br>(-0.37, 0.45) | 4.2%<br>(-30.9%, 56.9%) | 0.846 | -0.02<br>(-0.41, 0.38) | 1.7%<br>(-33.9%, 46.3%) | 0.934 |
| **Pressure**<br>**Injuries** | -0.09<br>(-0.56, 0.38) | -8.7%<br>(-43.0%, 46.3%) | 0.706 | -0.09<br>(-0.56, 0.38) | -8.3%<br>(-42.6%, 46.7%) | 0.719 |
| **Sepsis** | 0.10<br>(-0.17, 0.36) | 10.0%<br>(-15.7%, 43.6%) | 0.481 | 0.08<br>(-0.18, 0.35) | 8.6%<br>(-16.6%, 41.3%) | 0.540 |
| **CAUTI** | -0.18<br>(-0.32, -0.05) | -16.8%<br>(-27.2%, -<br>4.9%) | 0.007 | -0.19<br>(-0.32, -0.06) | -17.4%<br>(-27.6%, -<br>5.8%) | 0.004 |
| **CLABSI** | -0.04<br>(-0.21, 0.13) | -3.8%<br>(-18.8%,<br>13.9%) | 0.654 | -0.05<br>(-0.20, 0.11) | -4.4%<br>(-18.4%,<br>12.0%) | 0.578 |
| ***C. Diff*** | -0.002<br>(-0.12, 0.11) | -0.2%<br>(-10.9%,<br>11.8%) | 0.971 | -0.008<br>(-0.12, 0.10) | -0.8%<br>(-11.3%,<br>10.9%) | 0.891 |
| **MRSA** | -0.05<br>(-0.18, 0.08) | -4.7%<br>(-16.5%,<br>8.7%) | 0.474 | -0.06<br>(-0.19, 0.07) | -6.1%<br>(-17.4%,<br>6.8%) | 0.338 |

Log of the denominator of the outcome measure was included in the model as an off-set variable.
ADE, Adverse Drug Events; *C.diff*, *Clostridioides difficile*; CAUTI, catheter-associated urinary tract infection; CLABSI, central line-associated bloodstream infection; MRSA, Methicillin Resistant *Staphylococcus Aureus*

### Variables

*Outcome variables*: We assessed the impact of HQIC support on nine outcomes, including: 1) 30-day all-cause readmissions, 2) anticoagulant ADEs, 3) hypoglycemia ADEs, 4) pressure injuries, and 5) postoperative sepsis. The remaining four outcomes were HAIs: 6) CAUTI, 7) CLABSI, 8) C. diff, and 9) MRSA. Medicare Part A claims were used to calculate the numerator and denominator for the five non-HAI measures. Numerators for the four HAIs were obtained from NHSN and included all cases in the hospital, with the denominator including FFS beneficiaries discharged from each hospital. All nine outcomes were calculated at the hospital level by quarter. Details on the numerators and denominators for all outcome measures can be found in Table 3 in the Supplemental Material. In instances where the denominator was zero for an outcome (<0.5% of hospitals had missing data in a quarter), the quarterly data was excluded from the analyses.

### Covariates

The following hospital characteristics were considered as covariates for each model: hospital type (e.g., critical access hospital, tribal), hospital size measured through number of certified beds, rurality, and the level of socioeconomic disadvantage as measured by the Area Deprivation Index (ADI)^14^ of the hospital’s location. Characteristics of the patients in the outcome measures’ denominator were also included as covariates in the models for which these data were available (readmissions and ADEs models). These characteristics included percent of patients with a COVID-19 infection and percent of patients with specific demographic characteristics (e.g., age, gender, race and ethnicity, reason for Medicare eligibility) Table 4 in the Supplemental Material provides additional information on covariates.

### Analyses

We first conducted descriptive analyses to understand outcome distributions and the balance between the intervention and comparison groups. We matched each intervention hospital with a comparison hospital using exact matching for the following hospital characteristics: rural location, hospital type, hospital size, and ADI quantiles. We also applied propensity score matching,^15–17^ using caliper matching on the propensity score with a caliper size of 0.25. After matching, each comparison hospital was assigned a pseudo intervention start date based on the start date of their matched intervention hospital to allow each matched pair to have similar baseline and follow-up periods. Over 85% of hospitals in the intervention group were successfully matched with hospitals in the comparison group for most outcomes other than CAUTI (79.9% of hospitals), CLABSI (75.0% of hospitals) and sepsis (63.2% of hospitals). Sample sizes of the intervention groups for each of the outcomes before and after matching are in Supplemental Table 5.

To assess program effectiveness and determine the effect of receiving HQIC program support, we used a difference-in-difference (DiD) model specification with a Poisson generalized estimating equation (GEE) to compare the matched intervention and comparison hospitals. A DiD model was applied to each individual outcome to better isolate the impact of the HQIC program specifically. The Poisson GEE model is considered flexible in that it can handle repeated measurement of each subject (in this case, hospitals) and is robust against covariance misspecification. For each outcome, a log transformation was used to adjust the skewness of outcome data to better fit model assumptions.

The general structure of the DiD model used for each outcome is presented below:

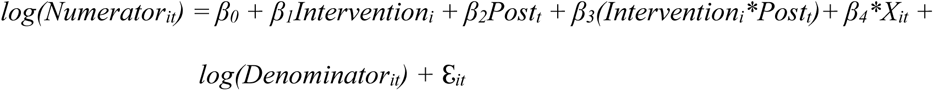

*Intervention* denotes whether hospital *i* received the HQIC program support for the outcome in quarter *t*. *Post* is the period indicator where 0 and 1 correspond to the baseline period and follow-up intervention period, respectively. The baseline period is the four quarters (i.e., one year) prior to a hospital beginning to receive HQIC support for the outcome. *X* represents hospital level covariates and *ℇ*represents the error term. The log of the denominator of the measures is included as an off-set variable. *β_3_* represents the DiD in the outcome rate between the intervention and non-intervention group comparing post and baseline periods, which is the main interest of the DiD model estimation. When a regression model did not converge, as was the case for outcomes with very low prevalence, covariates were systematically removed from the model until the model converged. In addition to the regression coefficients, we also report the associated relative rate of reduction calculated from the relative risk ratio between performance and baseline period.

## RESULTS

### Descriptive Statistics

Of the 1,929 hospitals enrolled in the program through December 2023, more hospitals (63.0%) received HQIC support on readmissions than other outcomes, followed by postoperative sepsis (56.9%). About half of hospitals received support for CAUTI (52.0%), C. diff (51.7%), CLABSI (51.4%), and MRSA (48.3%). Fewer hospitals received support for pressure injuries (44.4%), hypoglycemia ADEs (44.3%), and anticoagulant ADEs (42.6%).

Table 1 provides information about the hospitals included in the analytic samples for each outcome and the average baseline quarterly rates of the outcome measures, separated by intervention and comparison groups, after matching. The average baseline rate of hospital readmissions, pressure injuries, and CAUTI was significantly higher for hospitals in the intervention group than the rates of hospitals in the comparison group.

Tables 6-8 in the Supplemental Material show descriptive statistics of hospital characteristics for the intervention and comparison groups in each of the analytic samples.

### Intervention Activities

Figure 2 displays the proportion of hospitals receiving different types of intervention activities across the nine outcomes through December 31, 2023. Most hospitals (68% or higher) received training and technical assistance on evidence-based tools, skills-based training, and data analysis support. Fewer hospitals received support surrounding leadership engagement (42.7%), clinical workflow (32.8%), and technology (32.2%).

**Figure 2.**
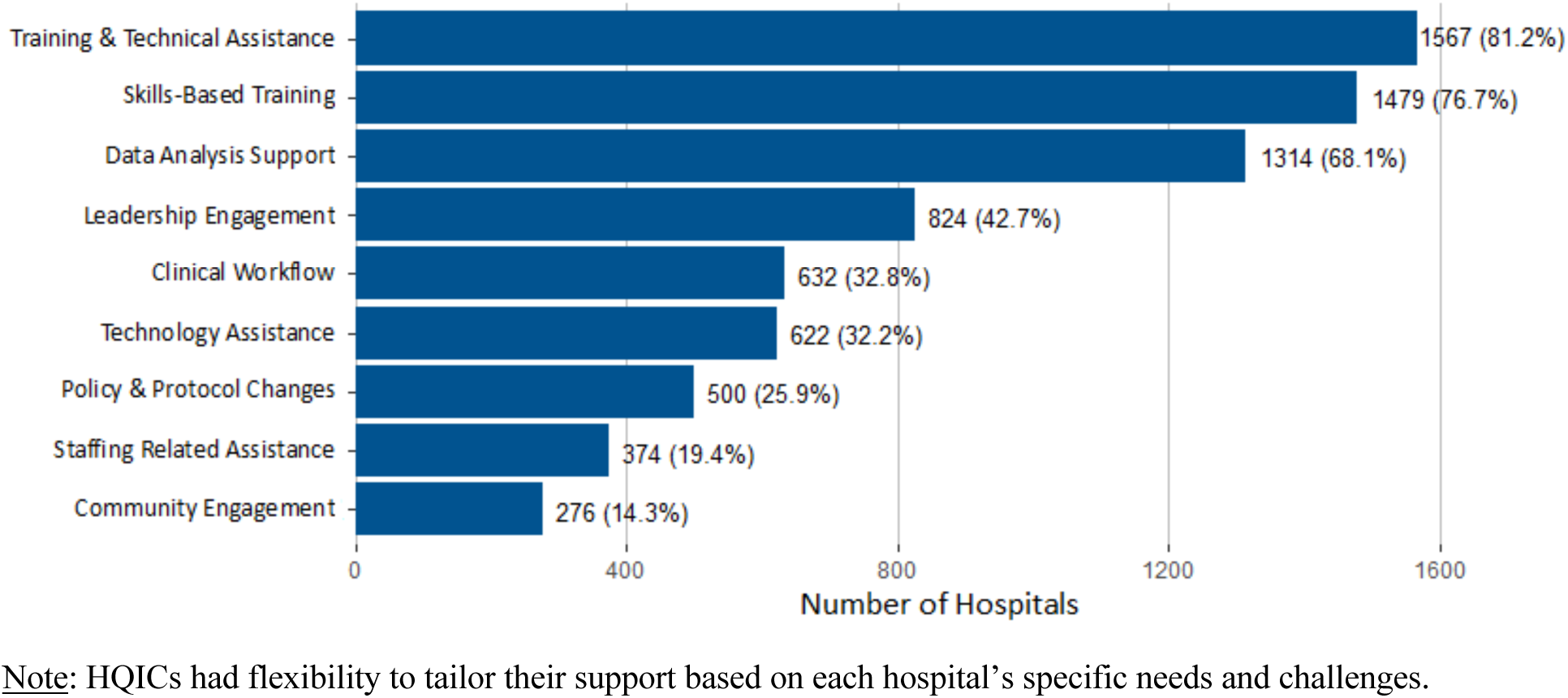
Type of Interventions Provided to Hospitals by Hospital Quality Improvement Contractors (HQICs) (N = 1,929 hospitals)

### Regression Analyses Results

Table 2 shows the results of the unadjusted and adjusted models estimating the effect of receiving HQIC program support on hospital readmissions and patient safety measures. In the unadjusted models, HQIC support was associated with a significant decrease in CAUTI events (coefficient = - 0.18, p <.01). After adjusting for covariates, results show that receipt of HQIC support resulted in significant reductions in 30-day all-cause readmissions (coefficient = -0.01, p <.05) and CAUTI events (coefficient = -0.19, p<.01). Receiving HQIC support was associated with a 1.4% reduction in the number of 30-day all-cause readmissions compared to hospitals with similar characteristics that did not receive HQIC support for readmissions. Hospitals receiving HQIC support for reducing CAUTI showed a 17.4% reduction in the number of CAUTI events relative to hospitals that did not receive support. There was no statistically significant effect of receiving HQIC program support on the other outcomes. Full model results can be found in Tables 9-17 in the Supplemental Material.

## DISCUSSION

HQIC support was associated with significant reductions in 30-day all-cause readmissions and CAUTI events. Both findings are congruent with the effort reported by HQICs on these outcomes. Although there was no statistically significant evidence of improvement in the other patient safety outcomes assessed, participating hospitals also saw nominal reductions in pressure injuries, MRSA, and CLABSI.

More enrolled hospitals received support for reducing readmissions than for any other outcome. The frequency at which readmissions occur and the fact that they are costly to healthcare settings likely made addressing readmissions a priority for HQICs and enrolled hospitals over other outcomes.

Hospital QI is an ongoing effort, and these results may reflect the experience and synergies created by several CMS QI initiatives. Additionally, the contractors are experienced with engaging hospital stakeholders and have a deep understanding of effective strategies for reducing readmissions. They used targeted interventions focusing on care transitions and incorporating best practices, such as enhanced care coordination, discharge planning, staff training, and patient education.^18^.

Readmissions carry a significant cost burden to the healthcare system and are a focus area for improvement for Medicare and policy makers.^9,19,20^ Even a small reduction in readmissions can have substantial cost savings. Using our model estimates, HQIC support resulted in an estimated 4,458 readmissions averted between October 1, 2021, and December 31, 2023. Based on an average cost of $12,426 per readmission (calculated using 2023 Medicare FFS claims), the associated cost savings is estimated to be $55.4M USD. Because the HQIC program did not provide hospitals with incentives or penalties, it is unlikely that the decrease in readmissions was a result of delays or decreases in needed admissions. Still, future analyses should examine this possibility.

CAUTIs are one of the most common HAIs and are associated with increased morbidity and mortality. The HQIC program was associated with a 17.4% reduction in CAUTI events. Reducing inappropriate urinary catheter use is considered one of the best ways to prevent CAUTI,^21^ and HQIC support likely influenced catheter use. In addition to the negative impact on patients, CAUTI treatment is costly, with an estimated cost of $15,807 per CAUTI event adjusted for 2021 inflation.^3^ Using the model estimates, approximately 524 CAUTI events were averted due to the program, resulting in $8.3M USD in cost savings.

Despite the impact on readmissions and CAUTI events, HQIC support was not associated with significant improvements across the remaining seven outcomes. These outcomes had very low baseline rates and in some cases (pressure injuries, adverse drug events, *C. diff*) have been decreasing nationwide^22,23^ providing hospitals with fewer opportunities to make notable improvements and/or making it more difficult to detect an impact. Over half of enrolled hospitals had 25 or fewer beds, and these outcomes were especially rare in those settings (<0.05% for most outcomes). Dedicating resources to reducing rare events may not have been possible for many hospitals or may not have been a priority. Although CAUTI events were also not common, a change needed to reduce CAUTI (i.e., reducing inappropriate catheter use) may have been easier to implement than some of the changes needed to reduce the other patient safety outcomes.

The COVID-19 public health emergency may have also limited the impact of the HQIC program. The contract period began in September 2020, a time when treating patients with COVID-19 and reducing risk of within-hospital COVID-19 infections was a top priority. The pandemic led to staffing shortages, turnover, and burnout.^24–26^ Staffing shortages have been a concern for rural hospitals and critical access hospitals for some time, and the pandemic may have worsened shortages,^27^ reducing focus on QI initiatives.

### Limitations

Several limitations should be noted. First, participation in the HQIC program was not randomly assigned, posing a threat to internal validity. The HQIC program deliberately supported hospitals that are small, in rural areas, and/or with lower quality ratings. Hospitals that accepted the support may have also differed in hospital priorities and motivation from those which did not. We addressed potential bias using a DiD approach, propensity score matching and by controlling for potential confounders. Even if not all differences were accounted for, the fact that hospitals perceiving the need for support improved in some outcomes over similar hospitals point to value for the program. The authors acknowledge that there are other overlapping programs that cover the same population with similar goals. These programs might have a spillover effect on QIO program impact estimation. We implemented a DiD model which assumes that other unobserved factors could affect both the intervention and comparison groups. The final DiD model estimation accounts for the impact of other programs.

Second, we relied on reported data from HQICs to determine which hospitals received support for a specific outcome. Assuming some degree of reporting error, some hospitals may have been misclassified in the intervention or comparison groups. Similarly, we could not confirm data on the number or types of intervention activities provided by HQICs or the fidelity of implementation of interventions by hospitals.

Third, we were not able to use the standardized infection ratio (SIR)^28^ for the HAI outcomes. This measure, calculated by the CDC and available in the NHSN, is used to track HAIs nationally and allows comparisons across facilities. The SIR measure is the ratio of observed events to the predicted number of events of the facility adjusting for facility- and patient-level factors that contribute to HAI risk. The predicted number of events was 0 for a large proportion of the hospitals in our samples.

Therefore, we used the number of events per hospital obtained from the NHSN as the numerator and the number of Medicare FFS discharges as the denominator. The rates we calculated are, thus, likely an overestimate of the true HAI rates, though this overestimation is unlikely to influence the findings on the impact of the program since it is unlikely that overestimation occurred more frequently in the intervention or the comparison hospitals.

Fourth, we could not adjust for all covariates in all the models due to the data not being available across all sources or due to the models not converging due to low prevalence of some of the outcomes. This omission may have introduced bias. When models did not converge, we dropped one variable at a time, trying to keep as many covariates as possible in the final model to help minimize the bias.

Finally, we were only able to examine HQIC program impact on Medicare FFS beneficiaries. The program could have had a different impact on outcomes for patients with other coverage.

## Conclusions

Our results show the impact of the HQIC program was mixed. Findings demonstrate HQIC support reduced readmissions and CAUTI but not the other targeted outcomes. For many of the hospitals targeted in this initiative, the support they received through the HQIC program on QI and data analytics was likely the only type of support they received in those areas. Many of the hospitals did not participate in other national quality of care monitoring^29^ or QI initiatives^19^ because of the small volume of patient stays. Our findings demonstrate that with support, these hospitals can make substantial improvement in quality of care. To further strengthen the impact of the program, stronger alignment between the outcomes targeted by the program and the prevalence of the outcomes in the targeted hospitals is needed, particularly for the smaller, rural hospitals that the HQIC program aims to support. Enhancing the measurement strategy could provide a better understanding of the program’s impact.

## Supporting information

All supplemental tables.

## Data Availability

No data are available. Although de-identified Medicare data can be requested through the Research Data Assistance Center (ResDAC) at https://resdac.org/ with a data use agreement, hospital identity, and data collected for the study are not available due to the quality improvement nature of the work.

