## Supplementary material for "Impact of Quality Improvement Support on Hospital Readmissions and Patient Safety Outcomes: A Quasi-Experimental Study of a National Quality Improvement Initiative": All supplemental tables.

### SUPPLEMENTAL MATERIAL

**Table 1. Description of Hospital Quality Improvement Contractor (HQIC) Intervention Activities**

| Intervention Activity | Description |
|---|---|
| Training and Technical Assistance on Evidence-Based Tools | Activities to directly support the implementation and adoption of evidence-based tools, such as checklists, educational handouts for patients and staff, risk assessment tools, action plans, and toolkits. |
| Skills-based Training | Development and/or implementation of training directed at hospital staff or representatives on evidence-based practices. |
| Data Analysis Support | Activities aimed at helping hospitals develop data collection protocols, develop analyses approaches, understand their data, and improve reporting capabilities to support quality improvement initiatives. |
| Leadership Engagement | Activities to directly engage leadership in hospital quality improvement efforts. |
| Technology Assistance | Activities to support hospitals in identifying, implementing, and improving technology solutions to increase efficiencies in data collection, hospital operations, and others. Examples of technology solutions include electronic health record checklists and dashboards that ensure appropriate data is collected and prompted. |
| Policy and Protocol Changes | Activities to develop, implement, and support the adoption of new or revised hospital policies and protocols to improve care coordination and reduce hospital acquired infections and adverse drug events. |
| Clinical Workflow | Activities to support the implementation, design, and adoption of new tools (e.g., checklists, flowcharts, electronic health record forms, action plans) to improve clinical workflow and hospital operations and efficiency. |
| Community Engagement | Activities to support hospitals in developing relationships with community-based organizations and integrating community resources to assist patients after discharge. Examples include community-based transportation services upon discharge, sharing information with patients about food banks/meals on wheels, hospitals working with pharmacies in the community to refer patients for diabetic testing if red flags arise. |
| Staffing-Related Assistance | Activities to help hospitals implement staff recruitment, onboarding, and retention, as well as training resources on quality improvement and other areas for new staff. |

**Table 2. Data Sources**

| Data Source | Description |
|---|---|
| CMS Hospital General Information File[a] | List of all hospitals registered with Medicare; the list includes addresses and hospital type. |
| Medicare Part A Claims | Main data used in analyses; used to assemble samples and to calculate outcome measures. |
| Medicare Beneficiary Information | Data includes demographics, enrollment, and eligibility information for Medicare beneficiaries. |
| HQIC Enrollment File | Monthly report of hospitals enrolled in HQIC program by CCN, enrollment information. |
| Intervention Template | Quarterly report by HQICs on interventions/technical assistance provided to their enrolled hospitals for each outcome. Used to identify intervention and comparison groups, and the intervention start date for intervention hospitals. |
| Area Deprivation Index (ADI)[b] | Measure of neighborhood socioeconomic disadvantage, created with census measures on income, education, poverty, employment, and housing quality. Ranges from 1-100, where the higher the number, the higher the level of disadvantage. Used the 2020 ADI to understand the level of socioeconomic disadvantage of the area in which the hospital is located. |
| CDC NHSN | A national HAI tracking system. This dataset was used to obtain the number of CAUTI, CLABSI, MRSA, and *C. diff* events per hospital each quarter for the numerators of the respective measures. |
| USDA Rural Urban Continuum Codes[c] | A classification system that distinguishes metropolitan counties by the population size of their metro area, and nonmetropolitan counties by degree of urbanization and adjacency to a metro area. Used to determine whether hospitals were located in a rural area. |

*Notes:*[a] Center for Medicare & Medicaid Services. Hospital General Information. Accessed February 2024. https://data.cms.gov/provider-data/dataset/xubh-q36u [b]Center for Health Disparities Research. Neighborhood Atlas: Area Deprivation Index (ADI) (2020 version). Accessed December 2023. https://www.neighborhoodatlas.medicine.wisc.edu [c] U.S. Department of Agriculture. Rural-Urban Continuum Codes (2023 version). Accessed date February 1, 2024 https://www.ers.usda.gov/data-products/rural-urban-continuum-codes.aspx

ADI, Area Deprivation Index; *C. diff, Clostridioides difficile*; CAUTI, catheter-associated urinary tract infection; CLABSI, central line-associated bloodstream infection; CCN, CMS Certification Number; CDC, Centers for Disease Control and Prevention; CMS, Centers for Medicare and Medicaid Services; HAI, Hospital Associated Infections; HQIC, Hospital Quality Improvement Contractor; MRSA, Methicillin Resistant Staphylococcus Aureus; NHSN, National Healthcare Safety Network; USDA, U.S. Department of Agriculture

**Table 3. Outcome Measures Specifications**

| Outcome | Numerator | Denominator |
|---|---|---|
| 30-day Hospital-wide, All-Cause Readmissions | Number of fees for service (FFS) inpatient discharges that have an inpatient admission to an acute care hospital or critical access hospital within 30-days of discharge. | Number of Medicare FFS inpatient discharges[a] at an acute care hospital or critical access hospital. Exclusions: discharges where beneficiaries were transferred to another hospital on the same day or one day following discharge; those discharged against medical advice; and discharges where, as of the discharge date, the beneficiary has passed away. |
| Anticoagulant ADEs | Number of FFS inpatient discharges with anticoagulant related ADEs not present on admission. ICD-10 Codes: T45511A, T45514A, T45515A | Number of Medicare FFS inpatient discharges.[a] |
| Hypoglycemia ADEs | Number of FFS inpatient discharges with hypoglycemic related ADEs, not present on admission. ICD-10 Codes: T383X1A, T383X4A, T383X5A | Number of Medicare FFS inpatient discharges.[a] |
| Pressure Injuries | Discharges among denominator, with any secondary ICD-10 diagnosis codes for pressure injury stage III or IV (or unstageable). Specifications can be found in AHRQ Quality Indicator (QI) specification (V2023, measure PSI 03)[b] | Number of Medicare FFS inpatient discharges for patients ages 18 years and older. Specifications for inclusion and exclusion criteria used are from the AHRQ QI, specification V2023, measure PSI 03.[b] |
| Post-Operative Sepsis | Discharges, among denominator, with any secondary ICD-10-CM diagnosis code for sepsis. Specifications can be found in AHRQ QI specification (V2023, measure PSI 13)[b] | Number of Medicare FFS inpatient discharges for elective surgeries for patients ages 18 years and older, with inpatient Medicare FFS stays. Specified inclusions can be found in AHRQ QI specification V2023, measure PSI 13.[b] |
| CAUTI | # of observed CAUTI events from all payers[c] | Number of Medicare FFS inpatient discharges.[a] |
| CLABSI | # of observed CLABSI events from all payers[c] | Number of Medicare FFS inpatient discharges.[a] |
| *C. diff* | # of observed *C. diff* events from all payers[c] | Number of Medicare FFS inpatient discharges.[a] |
| MRSA | # of observed MRSA events from all payers[c] | Number of Medicare FFS inpatient discharges.[a] |

*Notes:* [a]Using Medicare Part A claims. [b]Agency for Healthcare Research and Quality (AHRQ) Quality Indicator (QI): PSI Technical Specifications Updates. https://qualityindicators.ahrq.gov/measures/PSI_TechSpec [c]Data is obtained from the Centers for Disease Control and Prevention (CDC) National Healthcare Safety Network.

ADE, Adverse Drug Events; AHRQ QI, Agency for Healthcare Research Quality, Quality Indicators; *C. diff, Clostridioides difficile*; CAUTI, catheter-associated urinary tract infection; CLABSI, central line-associated bloodstream infection; FFS, Fee-For-Service; ICD-10, International Classification of Diseases ($10^{th}$ Revision); MRSA, Methicillin Resistant Staphylococcus Aureus; PSI, Patient Safety Indicators

**Table 4. Description of Covariates**

| Variables | Description | Data Source | Outcomes for which covariate was included in model |
|---|---|---|---|
| Hospital type | Type of hospital: Critical Access Hospital, Targeted IPPS, Urban IPPS, Rural IPPS, Tribal | Hospital General File | Readmission, CAUTI, CLABSI, *C. diff* |
| Hospital size | Based on number of certified inpatient beds, categorized as: 25 or less, 26-50, 51-100, 101-175, 176+ | HealthData.gov | Readmission, Anticoagulant ADE, Hypoglycemic ADE, Pressure Injury, Sepsis, CAUTI, MRSA, *C. diff* |
| Percentage of FFS patients in specified age groups | Percent of FFS patients in the denominator of the outcome measure in each of the following age groups: <65, 65-74, 75-84, 85+. Calculated quarterly for each outcome. | Medicare Beneficiary Information | Readmission, Anticoagulant ADE, Hypoglycemic ADE |
| Percent of FFS patients who are male | Percent of FFS patients in the denominator of the outcome measure with a hospital stay who were male. Calculated quarterly for each outcome. | Medicare Beneficiary Information | Readmission, Anticoagulant ADE, Hypoglycemic ADE |
| Percent of FFS patients from specified race and ethnicity groups | Percent of FFS patients with a hospital stay who were from each of the following racial and ethnic groups: White, Black, Asian/Pacific Islander, Hispanic, American Indian/Alaska Native, other/unknown race and ethnicity. Calculated quarterly for each outcome. | Medicare Beneficiary Information | Readmission, Anticoagulant ADE, Hypoglycemic ADE |
| Percent of FFS patients Medicare-eligible due to disability | Percent of FFS patients with a hospital stay who were eligible to Medicare due to disability. Calculated quarterly for each outcome. | Medicare Beneficiary Information | Readmission, Anticoagulant ADE, Hypoglycemic ADE |
| Percent of FFS dually eligible | Percent of FFS patients with a hospital stay who were eligible for Medicaid in addition to Medicare. Medicaid is a public insurance program for individuals with low income in the United States, funded jointly by the federal and state governments to provide free or low-cost healthcare services. Calculated quarterly for each outcome. | Medicare Beneficiary Information | Readmission, Anticoagulant ADE, Hypoglycemic ADE |
| Percent of FFS discharges with COVID-19 diagnosis | Percent of FFS patients with a hospital stay who had a COVID-19 infection as either a primary or secondary diagnosis ICD-10 diagnosis code in their claim. Calculated quarterly for each outcome. | Medicare Part A claims | Readmission, Anticoagulant ADE, Hypoglycemic ADE |

| Variables | Description | Data Source | Outcomes for which covariate was included in model |
|---|---|---|---|
| Rural location | Indicator variable of hospital's location being in a rural area. | USDA RUCC Codes[a] | Readmission, Anticoagulant ADE, Hypoglycemic ADE, Pressure Injury, Sepsis, CAUTI, CLABSI, MRSA, *C. diff* |
| ADI | Measure of the level of socioeconomic deprivation of the area in which the hospital is located. | Center for Health Disparities Research at the University of Wisconsin[b] | Readmission, Anticoagulant ADE, Hypoglycemic ADE, Pressure Injury, Sepsis, CAUTI, CLABSI, MRSA, *C. diff* |

[a] U.S. Department of Agriculture. Rural-Urban Continuum Codes (2023 version). Accessed date February 1, 2024 https://www.ers.usda.gov/data-products/rural-urban-continuum-codes.aspx [b] Center for Health Disparities Research. Neighborhood Atlas: Area Deprivation Index (ADI) (2020 version). Accessed December 2023. https://www.neighborhoodatlas.medicine.wisc.edu

ADE, Adverse Drug Events; ADI, Area Deprivation Index; *C. diff, Clostridioides difficile*; CAUTI, catheter-associated urinary tract infection; CLABSI, central line-associated bloodstream infection; IPPS, Inpatient Prospective Payment System (hospitals participating in Medicare's IPPS receive payment for hospital inpatient operating and capital-related costs based on a pre-determined rate for each hospital inpatient stay); MRSA, Methicillin Resistant Staphylococcus Aureus; USDA RUCC, U.S. Department of Agriculture Rural Urban Continuing Codes

**Table 5. Number of Hospitals in Intervention Group Before and After Matching by Outcome**

| Outcome | Hospitals in intervention group | Percent matched to a non-intervention (comparison) hospital (%) | Hospitals in intervention group included in analyses (N) |
|---|---|---|---|
| **Readmissions** | 1,206 | 85.9% | 1,036 |
| **Anticoagulant ADEs** | 814 | 90.4% | 736 |
| **Hypoglycemia ADEs** | 847 | 87.8% | 744 |
| **Pressure Injury** | 850 | 88.8% | 755 |
| **Sepsis** | 1,091 | 63.2% | 689 |
| **CAUTI** | 978 | 79.9% | 781 |
| **CLABSI** | 967 | 75.0% | 725 |
| ***C. diff*** | 910 | 86.5% | 787 |
| **MRSA** | 834 | 88.6% | 739 |

*Note:* Intervention groups include hospitals enrolled in the HQIC program that received HQIC support on the specific outcome by December 31, 2023.
ADE, Adverse Drug Event; *C. diff, Clostridioides difficile*; CAUTI, catheter-associated urinary tract infection; CLABSI, central line-associated bloodstream infection; MRSA, Methicillin Resistant Staphylococcus Aureus

**Table 6. Characteristics of matched samples: Readmissions, Anticoagulant ADEs, and Hypoglycemia ADEs**

| Characteristic | Readmissions Sample | | Anticoagulant ADEs Sample | | Hypoglycemia ADEs Sample | |
|---|---|---|---|---|---|---|
| | Intervention Group | Comparison Group | Intervention Group | Comparison Group | Intervention Group | Comparison Group |
| Number of Hospitals | 1,036 | 1,036 | 736 | 736 | 744 | 744 |
| ***Hospital Characteristics*** | | | | | | |
| Hospital Type | | | | | | |
| Critical Access Hospitals | 53.1% | 53.1% | 50.5% | 50.5% | 49.5% | 49.5% |
| Rural IPPS | 33.2% | 33.2% | 35.5% | 35.5% | 35.8% | 35.8% |
| Urban IPPS | 13.5% | 13.5% | 13.7% | 13.7% | 14.7% | 14.7% |
| Targeted IPPS & Tribal Hospitals | 0.2% | 0.2% | 0.3% | 0.3% | 0.1% | 0.1% |
| Hospital Size | | | | | | |
| 25 or Fewer Licensed Beds | 57.4% | 57.4% | 54.6% | 54.6% | 54.0% | 54.0% |
| 26-50 Licensed Beds | 14.5% | 14.5% | 15.2% | 15.2% | 14.5% | 14.5% |
| 51-100 Licensed Beds | 11.4% | 11.4% | 13.2% | 13.2% | 13.8% | 13.8% |
| 101-175 Licensed Beds | 7.0% | 7.0% | 7.9% | 7.9% | 7.8% | 7.8% |
| 176 or More Licensed Beds | 9.7% | 9.7% | 9.1% | 9.1% | 9.8% | 9.8% |
| Urban/Rural Location | | | | | | |
| Rural | 72.7% | 72.7% | 72.3% | 72.3% | 71.5% | 71.5% |
| Urban | 27.3% | 27.3% | 27.7% | 27.7% | 28.5% | 28.5% |
| Area Deprivation Index | 68.4 | 68.0 | 68.3 | 68.1 | 68.5 | 68.3 |
| ***Patient Characteristics*** | | | | | | |
| Age | | | | | | |
| % Under 65 | 12.0% | 11.3% | 11.7% | 12.3% | 12.0% | 12.1% |
| % 65-74 Years | 29.7% | 30.1% | 29.8% | 30.2% | 29.9% | 30.2% |
| % 75-84 Years | 32.7% | 32.7% | 32.9% | 32.6% | 32.6% | 32.8% |
| % 85+ Years | 25.5% | 25.5% | 25.4% | 25.0% | 25.2% | 24.7% |
| % Male | 43.4% | 43.4% | 44.0% | 44.4% | 44.1% | 44.2% |
| Race and Ethnicity | | | | | | |
| % Black | 5.7% | 5.4% | 5.1% | 6.2% | 5.5% | 6.3% |
| % White | 84.5% | 84.7% | 83.6% | 83.6% | 83.4% | 84.3% |
| % Hispanic | 5.2% | 4.0% | 6.4% | 4.0% | 6.3% | 3.8% |
| % Asian or Pacific Islander | 0.9% | 0.7% | 1.1% | 1.0% | 1.1% | 0.9% |
| % American Indian or Alaska Native | 1.3% | 2.2% | 1.5% | 2.1% | 1.3% | 1.6% |
| % Other/Unknown Race and Ethnicity | 1.3% | 1.4% | 1.4% | 1.5% | 1.4% | 1.4% |
| % Dually Eligible for Medicaid | 29.7% | 27.5% | 29.3% | 29.0% | 30.4% | 28.2% |
| % Medicare-Eligible due to Disability | 27.8% | 26.2% | 27.3% | 27.3% | 28.0% | 27.3% |

ADE, Adverse Drug Event; IPPS, Inpatient Prospective Payment System

**Table 7. Characteristics of matched samples: Pressure Injuries, Sepsis, CAUTI**

| Characteristic | Pressure Injuries Sample | | Sepsis Sample | | CAUTI Sample | |
|---|---|---|---|---|---|---|
| | Intervention Group | Comparison Group | Intervention Group | Comparison Group | Intervention Group | Comparison Group |
| Number of Hospitals | 755 | 755 | 689 | 689 | 781 | 781 |
| ***Hospital Characteristics*** | | | | | | |
| Hospital Type | | | | | | |
| Critical Access Hospitals | 49.1% | 49.0% | 40.1% | 39.9% | 44.5% | 44.5% |
| Rural IPPS | 35.6% | 35.8% | 42.7% | 43.0% | 41.2% | 41.2% |
| Urban IPPS | 15.1% | 15.0% | 17.2% | 17.1% | 14.3% | 14.3% |
| Targeted IPPS & Tribal Hospitals | 0.1% | 0.1% | 0.0% | 0.0% | 0.0% | 0.0% |
| Hospital Size | | | | | | |
| 25 or Fewer Licensed Beds | 53.5% | 53.3% | 45.5% | 45.3% | 49.2% | 49.4% |
| 26-50 Licensed Beds | 14.3% | 14.6% | 16.0% | 16.3% | 15.2% | 15.2% |
| 51-100 Licensed Beds | 12.9% | 12.8% | 15.5% | 15.5% | 15.9% | 15.7% |
| 101-175 Licensed Beds | 8.2% | 8.3% | 9.9% | 9.9% | 9.5% | 9.5% |
| 176 or More Licensed Beds | 11.0% | 11.1% | 13.1% | 13.1% | 10.2% | 10.2% |
| Urban/Rural Location | | | | | | |
| Rural | 71.2% | 71.4% | 68.5% | 68.7% | 70.4% | 70.4% |
| Urban | 28.8% | 28.6% | 31.5% | 31.3% | 29.6% | 29.6% |
| Area Deprivation Index | 67.5 | 67.7 | 65.4 | 65.2 | 67.3 | 67.2 |

ADE, Adverse Drug Event; CAUTI, catheter-associated urinary tract infection; IPPS, Inpatient Prospective Payment System

**Table 8. Characteristics of Matched Samples: CLABSI, *C. diff*, MRSA**

| Characteristic | CLABSI Sample | | *C. diff* Sample | | MRSA Sample | |
|---|---|---|---|---|---|---|
| | Intervention Group | Comparison Group | Intervention Group | Comparison Group | Intervention Group | Comparison Group |
| Number of Hospitals | 725 | 725 | 787 | 787 | 739 | 739 |
| ***Hospital Characteristic*****s** | | | | | | |
| Hospital Type | | | | | | |
| Critical Access Hospitals | 40.4% | 40.3% | 45.1% | 44.9% | 45.2% | 44.9% |
| Rural IPPS | 41.9% | 42.0% | 40.7% | 40.9% | 39.9% | 40.1% |
| Urban IPPS | 17.7% | 17.7% | 14.2% | 14.3% | 15.0% | 15.0% |
| Targeted IPPS & Tribal Hospitals | 0.0% | 0.0% | 0.0% | 0.0% | 0.0% | 0.0% |
| Hospital Size | | | | | | |
| 25 or Fewer Licensed Beds | 45.5% | 45.4% | 49.9% | 49.6% | 50.2% | 49.9% |
| 26-50 Licensed Beds | 15.2% | 15.2% | 15.5% | 15.6% | 16.2% | 16.3% |
| 51-100 Licensed Beds | 17.3% | 17.3% | 16.0% | 16.1% | 14.4% | 14.5% |
| 101-175 Licensed Beds | 9.1% | 9.1% | 8.7% | 8.7% | 8.7% | 8.8% |
| 176 or More Licensed Beds | 12.9% | 12.9% | 10.0% | 10.0% | 10.5% | 10.5% |
| Urban/Rural Location | | | | | | |
| Rural | 68.9% | 68.8% | 71.2% | 71.0% | 70.9% | 70.7% |
| Urban | 31.1% | 31.2% | 28.8% | 29.0% | 29.1% | 29.3% |
| Area Deprivation Index | 66.7 | 66.4 | 67.7 | 67.3 | 67.6 | 67.4 |

*C. diff, Clostridioides difficile*; CLABSI, central line-associated bloodstream infection; IPPS, Inpatient Prospective Payment System; MRSA, Methicillin Resistant Staphylococcus Aureus

**Table 9. Difference-in-Difference Regression Model: Readmissions (N = 27,082 hospital quarters)**

| Parameter | Estimate | 95% Confidence Limit | | P-value |
|---|---|---|---|---|
| Intercept | -2.283 | -2.374 | -2.192 | <0.0001 |
| Received Readmission Support | 0.019 | -0.0001 | 0.038 | 0.052 |
| Post Intervention | 0.003 | -0.006 | 0.012 | 0.512 |
| Post Intervention and Readmission Support | -0.014 | -0.026 | -0.002 | 0.028 |
| Rural (Reference: Urban) | -0.035 | -0.061 | -0.010 | 0.007 |
| Hospital Type (Reference: Urban IPPS) | | | | |
| Critical Access Hospital | -0.109 | -0.150 | -0.068 | <0.0001 |
| Rural IPPS | -0.076 | -0.107 | -0.045 | <0.0001 |
| Targeted IPPS & Tribal | 0.274 | -0.121 | 0.668 | 0.174 |
| Hospital Size (Reference: 25 or Fewer Licensed Beds) | | | | |
| 26-50 Licensed Beds | -0.020 | -0.060 | 0.020 | 0.332 |
| 51-100 Licensed Beds | 0.061 | 0.027 | 0.096 | 0.0005 |
| 101-175 Licensed Beds | 0.099 | 0.064 | 0.135 | <0.0001 |
| 176 or More Licensed Beds | 0.094 | 0.062 | 0.125 | <0.0001 |
| Percentage of Patients Ages < 65 | 0.120 | -0.050 | 0.290 | 0.166 |
| Percentage of Patients Ages 75 to 84 | 0.218 | 0.132 | 0.304 | <.0001 |
| Percentage of Patients Ages 85 or Older | 0.238 | 0.136 | 0.340 | <.0001 |
| Percentage of Male Patients | 0.076 | 0.004 | 0.148 | 0.038 |
| Percentage of Asian or Pacific Islander Patients | 0.293 | 0.090 | 0.496 | 0.005 |
| Percentage of Black Patients | 0.181 | 0.114 | 0.248 | <0.0001 |
| Percentage of Hispanic Patients | 0.064 | -0.009 | 0.138 | 0.087 |
| Percentage of American Indian/Alaska Native Patients | -0.081 | -0.249 | 0.087 | 0.345 |
| Percentage of Patients of Other[a]/Unknown Race and Ethnicity | 0.044 | -0.211 | 0.300 | 0.734 |
| Percentage of Patients Medicare-Eligible due to Disability | 0.378 | 0.289 | 0.466 | <0.0001 |
| Percentage of Patients Dually Eligible for Medicare and Medicaid | 0.316 | 0.251 | 0.380 | <0.0001 |
| Percentage of Patients with COVID-19 Diagnosis | 0.017 | -0.028 | 0.062 | 0.463 |
| Area Deprivation Index | 0.0008 | 0.0003 | 0.001 | 0.001 |

Notes: [a] Patient of other race and ethnicity refers to patients who are not White and who are not included in the other racial and ethnic categories.

Poisson generalized estimating equation. The outcome is log transformed. The log of the denominator of the outcome measure was included in the model as an offset variable.

IPPS, Inpatient Prospective Payment System

**Table 10. Difference-in-Difference Regression Model: Anticoagulant ADEs (N = 17,774 hospital quarters)**

| Parameter | Estimate | 95% Confidence Limit | | P-value |
|---|---|---|---|---|
| Intercept | -7.232 | -8.112 | -6.353 | <0.0001 |
| Received Anticoagulant Support | -0.042 | -0.247 | 0.164 | 0.691 |
| Post Intervention | -0.118 | -0.252 | 0.017 | 0.086 |
| Post Intervention and Anticoagulant Support | -0.026 | -0.197 | 0.146 | 0.770 |
| Rural (Reference: Urban) | -0.081 | -0.310 | 0.147 | 0.487 |
| Hospital Size (Reference: 25 or Fewer Licensed Beds) | | | | |
| 26-50 Licensed Beds | 0.268 | -0.094 | 0.629 | 0.146 |
| 51-100 Licensed Beds | 0.899 | 0.610 | 1.189 | <0.0001 |
| 101-175 Licensed Beds | 0.915 | 0.622 | 1.207 | <0.0001 |
| 176 or More Licensed Beds | 0.942 | 0.650 | 1.235 | <0.0001 |
| Percentage of Patients Ages < 65 | -2.753 | -4.404 | -1.101 | 0.001 |
| Percentage of Patients Ages 75 to 84 | -1.918 | -3.049 | -0.787 | 0.001 |
| Percentage of Patients Ages 85 or older | -0.528 | -1.711 | 0.656 | 0.382 |
| Percentage of Male Patients | 1.107 | 0.070 | 2.144 | 0.036 |
| Percentage of Asian or Pacific Islander Patients | 1.249 | -0.177 | 2.320 | 0.022 |
| Percentage of Black Patients | -0.631 | -1.428 | 0.166 | 0.121 |
| Percentage of Hispanic Patients | 0.092 | -0.715 | 0.900 | 0.823 |
| Percentage of American Indian/Alaska Native Patients | -0.396 | -2.972 | 2.180 | 0.763 |
| Percentage of Patients of Other[a]/Unknown Race and Ethnicity | -4.736 | -8.990 | -0.483 | 0.029 |
| Percentage of Patients Medicare-Eligible due to Disability | 1.031 | -0.269 | 2.231 | 0.120 |
| Percentage of Patients Dually Eligible for Medicare and Medicaid | -0.226 | -1.004 | 0.553 | 0.570 |
| Percentage of Patients with COVID-19 Diagnosis | 2.483 | 1.830 | 3.135 | <0.0001 |
| Area Deprivation Index | -0.009 | -0.014 | -0.003 | 0.002 |

Notes: [a] Patient of other race and ethnicity refers to patients who are not White and who are not included in the other racial and ethnic categories.
Poisson generalized estimating equation. The outcome is log transformed. The log of the denominator of the outcome measure was included in the model as an offset variable.
ADE, Adverse Drug Event

**Table 11. Difference-in-Difference Regression Model: Hypoglycemia ADEs (N = 18,675 hospital quarters)**

| Parameter | Estimate | 95% Confidence Limit | | P-value |
|---|---|---|---|---|
| Intercept | -10.388 | -12.669 | -8.107 | <0.0001 |
| Received Hypoglycemic Support | 0.005 | -0.353 | 0.363 | 0.978 |
| Post Intervention | -0.025 | -0.308 | 0.259 | 0.865 |
| Post Intervention and Hypoglycemic Support | 0.017 | -0.414 | 0.381 | 0.934 |
| Rural (Reference: Urban) | -0.282 | -0.727 | 0.163 | 0.214 |
| Hospital Size (Reference: 25 or Fewer Licensed Beds) | | | | |
| 26-50 Licensed Beds | 0.057 | -1.060 | 0.946 | 0.911 |
| 51-100 Licensed Beds | 0.239 | -0.373 | 0.850 | 0.444 |
| 101-175 Licensed Beds | 0.989 | 0.505 | 1.473 | <0.0001 |
| 176 or More Licensed Beds | 0.934 | 0.489 | 1.379 | <0.0001 |
| Percentage of Patients Ages < 65 | 0.079 | -4.487 | 4.644 | 0.973 |
| Percentage of Patients Ages 75 to 84 | -1.210 | -4.301 | 1.881 | 0.443 |
| Percentage of Patients Ages 85 or older | 0.058 | -3.130 | 3.246 | 0.972 |
| Percentage of Male Patients | 2.137 | -0.188 | 4.461 | 0.072 |
| Percentage of Asian or Pacific Islander Patients | 3.485 | 0.643 | 6.326 | 0.016 |
| Percentage of Black Patients | -0.540 | -1.837 | 0.757 | 0.414 |
| Percentage of Hispanic Patients | 0.415 | -0.932 | 1.761 | 0.546 |
| Percentage of American Indian/Alaska Native Patients | 0.729 | -2.286 | 3.744 | 0.636 |
| Percentage of Patients of Other[a]/Unknown Race and Ethnicity | -3.861 | -14.357 | 6.635 | 0.471 |
| Percentage of patients Medicare-Eligible due to Disability | 0.967 | -2.940 | 4.873 | 0.628 |
| Percentage of Patients Dually Eligible for Medicare and Medicaid | -0.910 | -2.390 | 0.570 | 0.228 |
| Percentage of Patients with COVID-19 Diagnosis | 0.704 | -0.966 | 2.374 | 0.409 |
| Area Deprivation Index | -0.003 | -0.011 | 0.005 | 0.430 |

Notes: [a] Patient of other race and ethnicity refers to patients who are not White and who are not included in the other racial and ethnic categories.
Poisson generalized estimating equation. The outcome is log transformed. The log of the denominator of the outcome measure was included in the model as an offset variable.
ADE, Adverse Drug Event

**Table 12. Difference-in-Difference Regression Model Estimates: Pressure Injury (N = 18,369 hospital quarters)**

| Parameter | Estimate | 95% Confidence Limit | | P-value |
|---|---|---|---|---|
| Intercept | -9.288 | -9.906 | -8.671 | <0.0001 |
| Received Pressure Injury Support | 0.527 | 0.177 | 0.877 | 0.003 |
| Post Intervention | -0.265 | -0.637 | 0.108 | 0.164 |
| Post Intervention and Pressure Injury Support | -0.086 | -0.556 | 0.383 | 0.719 |
| Hospital Size (Reference: 25 or Fewer Licensed Beds) | | | | |
| 26-50 Licensed Beds | -0.161 | -1.080 | 0.758 | 0.731 |
| 51-100 Licensed Beds | 0.520 | -0.074 | 1.115 | 0.086 |
| 101-175 Licensed Beds | 1.077 | 0.459 | 1.695 | 0.0006 |
| 176 or More Licensed Beds | 1.131 | 0.629 | 1.640 | <0.0001 |
| Area Deprivation Index | -0.003 | -0.008 | 0.003 | 0.301 |

Poisson generalized estimating equation. The outcome is log transformed. The log of the denominator of the outcome measure was included in the model as an offset variable.

**Table 13. Difference-in-Difference Regression Model Estimates: Postoperative Sepsis (N = 11,344 hospital quarters)**

| Parameter | Estimate | 95% Confidence Limit | | P-value |
|---|---|---|---|---|
| Intercept | -5.612 | -5.932 | -5.293 | <0.0001 |
| Received Sepsis Support | 0.220 | 0.002 | 0.438 | 0.048 |
| Post Intervention | -0.040 | -0.215 | 0.134 | 0.650 |
| Post Intervention and Sepsis Support | 0.082 | -0.181 | 0.346 | 0.540 |
| Rural (Reference: Urban) | 0.008 | -0.417 | 0.433 | 0.970 |
| Hospital Type (Reference: Urban IPPS) | | | | |
| Critical Access Hospital | -1.501 | -2.246 | -0.756 | <0.0001 |
| Rural IPPS | -0.104 | -0.556 | 0.349 | 0.653 |
| Hospital Size (Reference: 25 or Fewer Licensed Beds) | | | | |
| 26-50 Licensed Beds | -0.348 | -0.838 | 0.142 | 0.163 |
| 51-100 Licensed Beds | 0.146 | -0.342 | 0.634 | 0.558 |
| 101-175 Licensed Beds | 0.309 | -0.070 | 0.688 | 0.110 |
| 176 or More Licensed Beds | 0.371 | 0.073 | 0.670 | 0.015 |
| Area Deprivation Index | -0.001 | -0.006 | 0.003 | 0.574 |

Poisson generalized estimating equation. The outcome is log transformed. Targeted IPPS & Tribal category is excluded from the hospital type due to no records associated with this rare event measure. The log of the denominator of the outcome measure was included in the model as an offset variable.
IPPS, Inpatient Prospective Payment System

**Table 14. Difference-in-Difference Regression Model Estimates: CAUTI (N = 17,839 hospital quarters)**

| Parameter | Estimate | 95% Confidence Limit | | P-value |
|---|---|---|---|---|
| Intercept | -5.930 | -6.260 | -5.601 | <0.0001 |
| Received CAUTI Support | 0.313 | 0.118 | 0.509 | 0.002 |
| Post Intervention | -0.210 | -0.319 | -0.101 | 0.0002 |
| Post Intervention and CAUTI | -0.191 | -0.323 | -0.060 | 0.004 |
| Rural (Reference: Urban) | -0.050 | -0.260 | 0.160 | 0.6411 |
| Hospital Type (Reference: Urban IPPS) | | | | |
| Critical Access Hospital | -0.724 | -1.077 | -0.372 | <0.0001 |
| Rural IPPS | -0.432 | -0.673 | -0.191 | 0.0004 |
| Hospital Size (Reference: 25 or Fewer Licensed Beds) | | | | |
| 26-50 Licensed Beds | -0.039 | -0.420 | 0.342 | 0.841 |
| 51-100 Licensed Beds | 0.074 | -0.345 | 0.494 | 0.729 |
| 101-175 Licensed Beds | 0.094 | -0.251 | 0.440 | 0.592 |
| 176 or More Licensed Beds | 0.205 | -0.100 | 0.511 | 0.187 |
| Area Deprivation Index | -0.002 | -0.008 | 0.004 | 0.502 |

Poisson generalized estimating equation. The outcome is log transformed. Targeted IPPS & Tribal category is excluded from the hospital type due to no records associated with this rare event measure. The log of the denominator of the outcome measure was included in the model as an offset variable.
CAUTI, catheter-associated urinary tract infection; IPPS, Inpatient Prospective Payment System

**Table 15. Difference-in-Difference Regression Model Estimates: CLABSI (N = 16,284 hospital quarters)**

| Parameter | Estimate | 95% Confidence Limit | | P-value |
|---|---|---|---|---|
| Intercept | -6.132 | -6.477 | -5.787 | <0.0001 |
| Received CLABSI Support | 0.228 | 0.018 | 0.437 | 0.033 |
| Post Intervention | -0.222 | -0.341 | -0.103 | 0.0003 |
| Post Intervention and CLABSI Support | -0.045 | -0.203 | 0.113 | 0.578 |
| Rural (Reference: Urban) | 0.050 | -0.207 | 0.307 | 0.702 |
| Hospital Type (Reference: Urban IPPS) | | | | |
| Critical Access Hospital | -2.924 | -3.459 | -2.390 | <0.0001 |
| Rural IPPS | -0.902 | -1.209 | -0.595 | <0.0001 |
| Area Deprivation Index | 0.008 | 0.003 | 0.013 | 0.002 |

Poisson generalized estimating equation. The outcome is log transformed. Targeted IPPS & Tribal category is excluded from the hospital type due to no records associated with this rare event measure. The log of the denominator of the outcome measure was included in the model as an offset variable.
CLABSI, central line-associated bloodstream tract infection; IPPS, Inpatient Prospective Payment System

**Table 16. Difference-in-Difference Regression Model Estimates: *C. diff* (N = 18,348 hospital quarters)**

| Parameter | Estimate | 95% Confidence Limit | | P-value |
|---|---|---|---|---|
| Intercept | -5.162 | -5.397 | -4.927 | <0.0001 |
| Received *C. diff* Support | 0.157 | 0.0003 | 0.314 | 0.0496 |
| Post Intervention | -0.119 | -0.203 | -0.035 | 0.0057 |
| Post Intervention and *C. diff* Support | -0.008 | -0.120 | 0.104 | 0.8908 |
| Rural (Reference: Urban) | 0.029 | -0.170 | 0.229 | 0.7744 |
| Hospital Type (Reference: Urban IPPS) | | | | |
| Critical Access Hospital | -0.322 | -0.614 | -0.030 | 0.031 |
| Rural IPPS | -0.287 | -0.514 | -0.060 | 0.013 |
| Hospital Size (Reference: 25 or Fewer Licensed Beds) | | | | |
| 26-50 Licensed Beds | 0.321 | 0.082 | 0.560 | 0.009 |
| 51-100 Licensed Beds | 0.409 | 0.160 | 0.658 | 0.001 |
| 101-175 Licensed Beds | 0.197 | -0.133 | 0.526 | 0.242 |
| 176 or More Licensed Beds | 0.225 | -0.050 | 0.499 | 0.109 |
| Area Deprivation Index | -0.005 | -0.009 | -0.0001 | 0.044 |

Poisson generalized estimating equation. The outcome is log transformed. Targeted IPPS & Tribal category is excluded from the hospital type due to no record associated with this rare event measure. Log of the denominator of the outcome measure was included in the model as an offset variable.
*C. diff, Clostridioides difficile*; IPPS, Inpatient Prospective Payment System

**Table 17. Difference-in-Difference Regression Model Estimates: for MRSA (N = 17,239 hospital quarters)**

| Parameter | Estimate | 95% Confidence Limit | | P-value |
|---|---|---|---|---|
| Intercept | -6.674 | -6.916 | -6.431 | <0.0001 |
| Received MRSA | 0.290 | 0.090 | 0.490 | 0.005 |
| Post Intervention | -0.182 | -0.271 | -0.093 | <0.0001 |
| Post Intervention and MRSA Support | -0.063 | -0.191 | 0.066 | 0.338 |
| Rural (Reference: Urban) | -0.036 | -0.319 | 0.246 | 0.801 |
| Hospital Type (Reference: Urban IPPS) | | | | |
| Critical Access Hospital | -1.719 | -2.147 | -1.290 | <0.0001 |
| Rural IPPS | -0.655 | -0.946 | -0.364 | <0.0001 |
| Hospital Size (Reference: 25 or Fewer Licensed Beds) | | | | |
| 26-50 Licensed Beds | 0.133 | -0.177 | 0.443 | 0.401 |
| 51-100 Licensed Beds | 0.472 | 0.180 | 0.767 | 0.002 |
| 101-175 Licensed Beds | -0.081 | -0.453 | 0.291 | 0.668 |
| 176 or More Licensed Beds | 0.013 | -0.325 | 0.350 | 0.940 |
| Area Deprivation Index | 0.002 | -0.003 | 0.007 | 0.386 |

Poisson generalized estimating equation. The outcome is log transformed. Targeted IPPS & Tribal category is excluded from the hospital type due to no record associated with this rare event measure. The log of the denominator of the outcome measure was included in the model as an offset variable.
IPPS, Inpatient Prospective Payment System; MRSA, Methicillin Resistant Staphylococcus Aureus
